# The development of the gynecologic oncology pathway “VivaPathway GT“ – a qualitative study about the transformation from an implicit to an explicit, evidence-based clinical pathway in a Berlin-based tertiary care hospital

**DOI:** 10.1101/2024.04.14.24305788

**Authors:** Anne Büscher, Nicolle Reinhold, Joachim Kugler

## Abstract

**Introduction:** Standardization in healthcare is crucial for comprehensive patient care, as emphasized by the WHO and quality management principles. Clinical pathways offer a structured approach to describing essential processes, particularly relevant in gynecologic oncology care. Despite their proven benefits, pathways remain underutilized, highlighting the need for explicit translation of implicit pathways.

**Materials and Methods:** This study employed semi-structured interviews with healthcare professionals to capture implicit gynecologic oncology pathway. Interviews were examined using qualitative content analysis. External requirements, legal mandates, and certification criteria were integrated into pathway development. A participatory approach involving interprofessional collaboration guided pathway refinement.

**Results:** The study applied the Pathway Association’s 7-phase method, illustrating the development of VivaPathway GT. The explicit pathway, enriched with external requirements, comprised 26 steps, enhancing comprehensiveness and interprofessional involvement. Notable additions included pre-therapeutic tumor boards and detailed post-operative consultations.

**Discussion:** Transitioning from implicit to explicit pathways is a is a demanding and challenging process emphasizing stakeholder engagement and IT support. Financial constraints pose challenges, but initiatives like quality contracts offer opportunities for resource allocation. The study validates hypotheses, confirming the efficacy of collaboration of involved professionals and integration of external requirements in pathway development.

**Conclusions:** This study underscores the importance of explicit clinical pathways. It provides valuable insights for healthcare professionals facing similar challenges, emphasizing the need for ongoing evaluation and adaptation. Despite resource challenges, stakeholder collaboration and IT support are crucial for pathway effectiveness and relevance in evolving healthcare landscapes.

## Introduction

Standardization in healthcare is a central factor in enhancing comprehensive patient care, emphasized by a clear call from the WHO [1]. In the healthcare management context, this entails describing processes within the domain of process quality. It represents the second dimension of quality management, alongside the first dimension of structural and the third dimension of outcome quality [2].

The patient, with their individual journey, is at the center of inpatient healthcare. This directs healthcare professionals in describing the essential processes through the implementation of clinical pathways. In a Cochrane review Rotter et al. [3] demonstrate that the use of clinical pathways has positive effects on the following outcomes: reduced likelihood of in-hospital complications, improved documentation quality, shorter length of stay, and reduced hospital costs. No differences were observed in terms of hospital readmission and in-hospital mortality. Moreover, the Diagnosis-Related Group (DRG) system is clearly associated with the standardization of these processes, requiring them for structured resource management [4]. Furthermore, in pursuit of the mitigation of liability risks, hospitals are advised to implement and rely on precise standardized processes [5].

In the future, the worth of standardizing clinical processes will continue to grow. Political directives in Germany already actively demand such standardization in other clinical areas. For instance, in the Guideline of the Joint Federal Committee (G-BA) on Measures for Quality Assurance in the Care of Patients with Proximal Femoral Fracture (QSFFx-RL) guideline, seven explicit Standard Operating Procedures (SOPs) are required [6]. The state government of Berlin designates certified centers with specific tasks (e. g. oncological centers) in hospital planning [7]. To meet these requirements, clinical pathways are an integral component. A recent study by Schmitt et al. [8] provides evidence of enhanced patient care in certified oncology clinics. The results show advantages in overall survival rates if cancer patients are initially treated in certified cancer centers. Thus, one of the largest hospitals in Berlin, a tertiary care hospital, is actively preparing for the certification as an oncological center.

The future calls for an increased emphasis on standardizing clinical processes to effectively address the diverse demands mentioned and create positive outcomes for all stakeholders. Rotter et al. [9] described the three main characteristics of clinical pathways. The first quality is the translation of evidence-based medicine into existing (hospital) structures. Secondly, pathways outline a detailed sequence of steps. Thirdly, it describes the scope for a specific clinical issue or procedure.

The implementation of a standardized clinical pathways entails an organizational change. It is necessary to recognize that achieving successful change requires more than the implementation of standardized pathways; active engagement from both clinical and management staff is needed. According to Rotter et al. [9] “Passive, top-down strategies to promote and implement CPWs [Clinical Pathways] have little or no impact. Engagement of both clinical and management staff in the development and adoption of CPWs is required and multifaceted strategies should be used to implement this concept.”

Unfortunately, the valuable and necessary tool of CPWs is underutilized. In 2006, a global survey by the European Pathway Association found limited adoption of clinical pathways. Highest rates were in Estonia, Singapore, and the United States. Germany, along with other countries, reported usage for only 6 – 10 % of patients [10]. This suggests that if standards in clinical settings are followed, they are mainly implicitly adhered to. Thus, the term implicit pathway refers to the pathway that is lived in practice but not formally documented. These informal pathways represent the standards according to which staff routinely operate, based on their experiences and internal guidelines, without being explicitly documented. Explicit pathways are formally documented. In the mentioned tertiary care hospital, there is currently no explicit clinical pathway for gynecologic oncology care.

However, to avoid uncertainties and errors, meet the requirements and to empower all benefits of institutional knowledge management, implicitly pathways must be translated into explicit ones.

It leads to the following research question of this study: How can an implicit pathway be translated into an evidence-based, explicit clinical pathway? By using the example of a gynecologic oncology clinical pathway in a tertiary care hospital in Berlin.

The change management concept according to Lewin [11] will serve as a theoretical basis. He describes that participatory management approach involves active employee engagement in decision-making processes and the design of their work environment. It aims to recognize employees as valuable sources of ideas, experiences, and suggestions and encourages their involvement in improving work processes.

Based on this participatory approach the following hypotheses are formulated.

1. The translation of implicit clinical pathways into an explicit one through an interprofessional approach can be facilitated by integrating insights from interviews with involved health professionals in collaborative processes.
2. Adding external requirements, especially legal mandates, during the pathway development creates the essential connection between external demands and internal structures, cultivating a holistic and evidence-based explicit pathway.

## Material and Methods

In order to address the research question and test the formulated hypotheses, semi-structured interviews will be conducted with involved health professionals. Thus, an interprofessional approach at the development process of the explicit clinical pathway will be used. These data will be analyzed using qualitative content analysis following Mayring’s methodology [12]. The implicit gynecological oncological pathway will be captured through inductive coding. To enhance quality, the analysis will be independently performed by the second author.

In the second phase, guidelines and certification criteria, and legal requirements will be integrated into the coded sequence. Both the first and second authors contribute to this process.

Subsequently, a quality circle is convened, involving all stakeholders, including quality and process management. The implicit pathway, now enriched with external requirements, serves as the basis for discussion. Additional steps are integrated based on the preferences of health professionals.

### Ethical Considerations in non-interventional research with healthcare professionals

The design of this research - a study involving semi-structured interviews with healthcare professionals - does not require an ethical approval. The study focuses on gathering insights into professional practices and perspectives. All participants were informed about the study’s objectives and their rights, with written consent obtained for participation and audio recording, ensuring voluntary engagement and comprehension of the research’s aims. Furthermore, as the participants are healthcare professionals familiar with ethical standards, the shared information was based on professional expertise, distinguishing our study from those potentially impacting patients or the public.

### Selection of interview participants

A total of 20 participants were interviewed. These participants not only represent the main stakeholders of the gynecologic oncology pathway but also include all other professional groups involved in this clinical and management process. The interview participants are four gynecologists, as well as one physician from the specialties of oncology, urology, palliative medicine, radiology, visceral surgery, pathology, radiation therapy, nuclear medicine, and anesthesia/pain medicine. All interviewed physicians held the position of chief or senior physician in their respective departments at the mentioned tertiary care hospital and have at least ten years of clinical experience. All of them are already involved in the implicitly used clinical pathway of gynecological oncological patients.

Furthermore, one representative from the following professional groups, each with at least five years of professional experience, was interviewed: psycho-oncology, nutritional science, physiotherapy, social services, nursing care of oncological unit, nursing admission management, and tumor documentation.

The interview participants were jointly selected by the first and the second authors. The second author is the head senior physician of the gynecologic tumor surgery department, while the first author is involved in hospital management. The interview participants were chosen to ensure representation of all professional groups currently involved in the care of gynecologic oncology patients. Additionally, the certification criteria and corresponding evidence-based guidelines were reviewed to determine if there were any other possible professional groups to include. However, all necessary professional groups are already involved in the process of gynecological oncological patients. The interviews were conducted on a voluntary basis; no requested interviewee declined to participate in the study.

### Data collection – Interviews

The recruitment period for this study began on May 20, 2022, and ended on March 13, 2023. In July and August 2022, the interviews were conducted by the first author. A final interview took place in March 2023. The majority of the interviews (13), were conducted via a video conferencing platform, while seven were conducted face-to-face. The latter took place in offices at the tertiary care hospital in Berlin. All interviews were carried out in the native language of all participants, which is German, and were recorded with the consent of the interviewees using a voice recorder app. The duration of the interviews ranged from 15 to 50 minutes, with a median duration of 30 minutes. Apart from the interviewee and the interviewer, there were no other participants or observers.

All interviews were semi-structured. The interview participants were divided into two groups. Firstly, the group of the four gynecological physicians. These participants are from the leading department and, therefore, accompany patients from admission to discharge. Secondly, all other interviewees involved in individual steps of the clinical process. Consequently, two different questionnaires were developed to explore the current implicit pathway.

To develop the first interview questionnaire, the second author, the head senior physician of the gynecological department, was consulted regarding the current implicit pathway. In order to develop a questionnaire, questions were asked along this implicit process with 14 steps. At each step of these steps, several questions were developed to ask during the following interviews. About 1. how this step is reached, 2. what the content of this step is, and if 3. there are any standards like SOPs. In addition, it is asked what and 4. where it is documented, and 5. how the further process proceeds after this step. This questionnaire was tested with the head senior physician and adjusted accordingly. Thus, a structure and sequence of questions were established. However, there was sufficient flexibility to provide feedback on missing steps at any time. All gynecologists were interviewed using this first questionnaire. All other professional groups were interviewed using a second questionnaire. It starts by inquiring at 1. which step in the pathway they are contacted and 2. how. Furthermore, they are asked 3. if they are contacted again in the patient’s process. Additionally, it was investigated 4. who or what determines whether their involvement follows. If so, 5. what purpose is achieved through their work and 6. where it is documented. This second questionnaire was also tested with one interviewee and then slightly adjusted. In addition, all interview participants were asked for any suggestions to improve both patient care and employee satisfaction. This request was made to ensure continuous improvement and engagement of all stakeholders in the development of new standards. The goal is to uphold the quality of care, especially with the anticipated increase in the number of patients in the future. At the beginning of the interviews, the participants were informed about the purpose of the survey, and all gave their consent for data processing.

### Interview Analysis

The transcription of interview recordings was performed using the automated transcription service Amberscript. To enhance quality, the first author corrected the transcripts where necessary. The Mayring content analysis method was used [12].

An inductive approach was applied, as there were no existing codes that could serve as a basis. Even though the implicit process described by the senior physician (2. Author) formed the structure for the questionnaire, there was sufficient flexibility in the application of both questionnaires to recognize new process steps, or codes. These new codes could then serve as a basis in developing the explicit pathway. The transcripts were initially divided into parts; firstly, the results of gynecologists and secondly the answers of all other professionals. These sections were then paraphrased, summarized, and developed for category formation. Excel was used as the basis for this process.

The interviews with the gynecologists were analyzed in this manner by the first author, and then the codes were adjusted. The second author also analyzed the interviews for category formation. Different interpretations were discussed, and a common solution for a code was found.

The codes were named after the various steps of the implicit clinical pathway for gynecological oncological patients. Subsequently, the quality circle was conducted. All interview participants were invited. In a participatory approach here, the final explicit pathway was finalized and described in a digital flowchart.

The qualitative research methodology was conducted in accordance with the Quality Assurance Criteria for Reporting Qualitative Research (COREQ) checklist. [13].

### Synthesizing Research Findings and Applying a 7-Phase Method for CPW Development

These research findings can provide support to others planning similar initiatives when the approach is presented in a concise and clear overview. Therefore, an existing theoretical framework will be utilized in a subsequent step. The Pathway Association has developed a 7-phase method for clinical pathways which assist in the development, implementation, and evaluation of clinical pathways [14]. The steps described above will be categorized into these phases, resulting in a brief overview of the process.

## Results

The results section is presented in three parts. The first part demonstrates the application of the theoretical framework by the Pathway Association (table 2). In this section, the approach to developing the pathway is described. The second part illustrates the VivaPathway GT (flowchart 1), demonstrating the results of the interviews and additions of external requirements. The results section concludes with the third part. This section provides an overview of the differences between implicit and explicit pathways, emphasizing the changes from previously unwritten standards to the now finalized and formal pathway.

**Table 1:** Characteristics of interview participants / Numbers of persons.

|  |  |  |  |  |  |  |  |  |  |  |
| --- | --- | --- | --- | --- | --- | --- | --- | --- | --- | --- |
| Interview-participants<br>Medical doctors<br>> 10 yrs of clinical experiences | Gyne-cology | Oncology | Urology | Palliative medicine | Radiology | Visceral surgery | Pathology | Radiation therapy | Anesthesia / pain medicine | Nuclear medicine |
|  | 4 | 1 | 1 | 1 | 1 | 1 | 1 | 1 | 1 | 1 |
| Interview-participants<br>Other involved professionals<br>> 5 yrs of professional experience | Psycho-oncology | Nutritional science | Physiotherapy | Social services |  | Nursing care of oncological unit |  | Nursing admission management | Tumor documentation |  |
|  | 1 | 1 | 1 | 1 |  | 1 |  | 1 | 1 |  |
| Age group | 30 – 39 |  |  | 40 – 49 |  |  | 50 – 59 |  |  | 60 -69 |
|  | 4 |  |  | 6 |  |  | 7 |  |  | 3 |
| Gender | Female |  |  |  |  | Male |  |  |  |  |
|  | 10 |  |  |  |  | 10 |  |  |  |  |

**Table 2:**
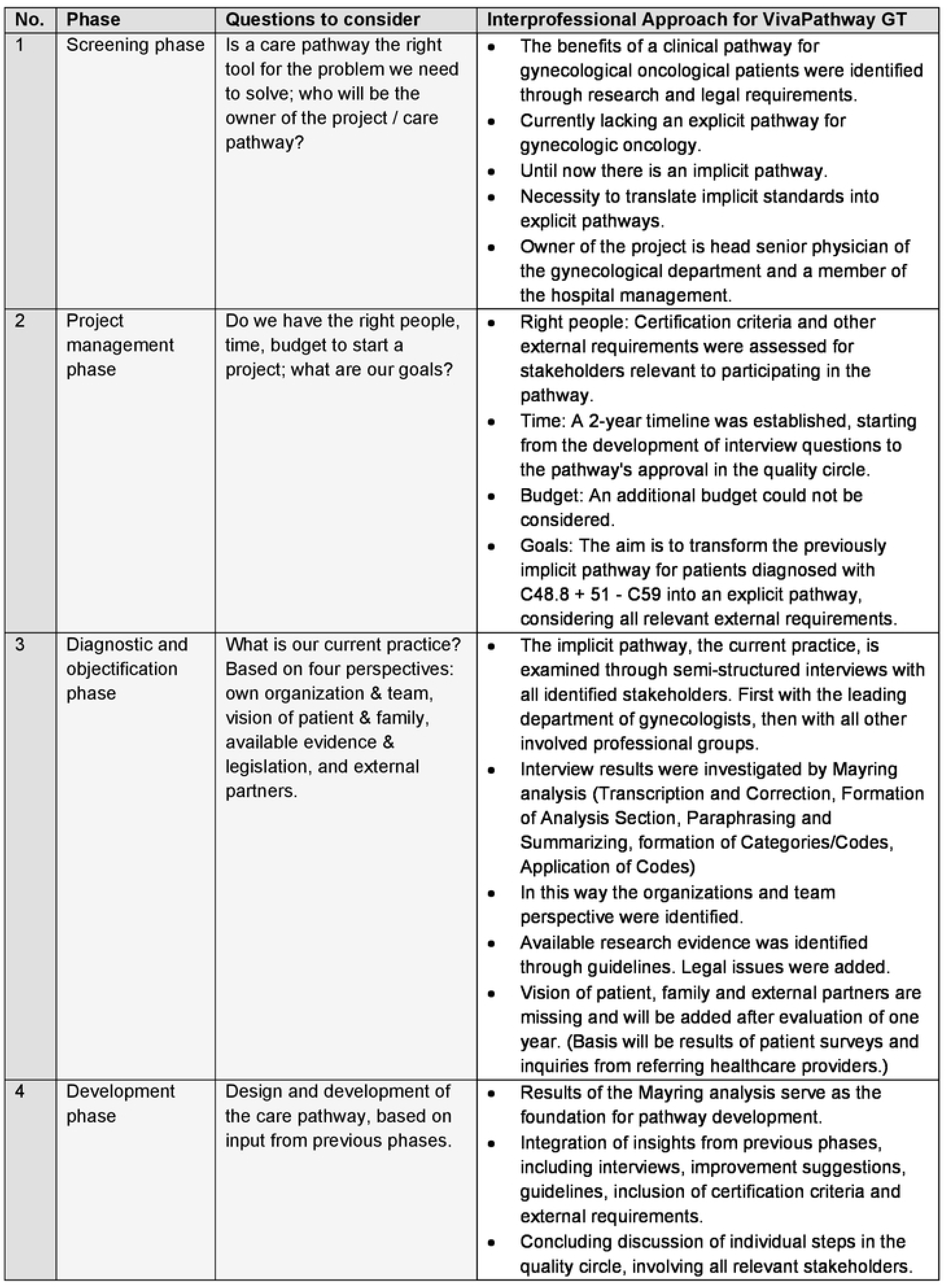
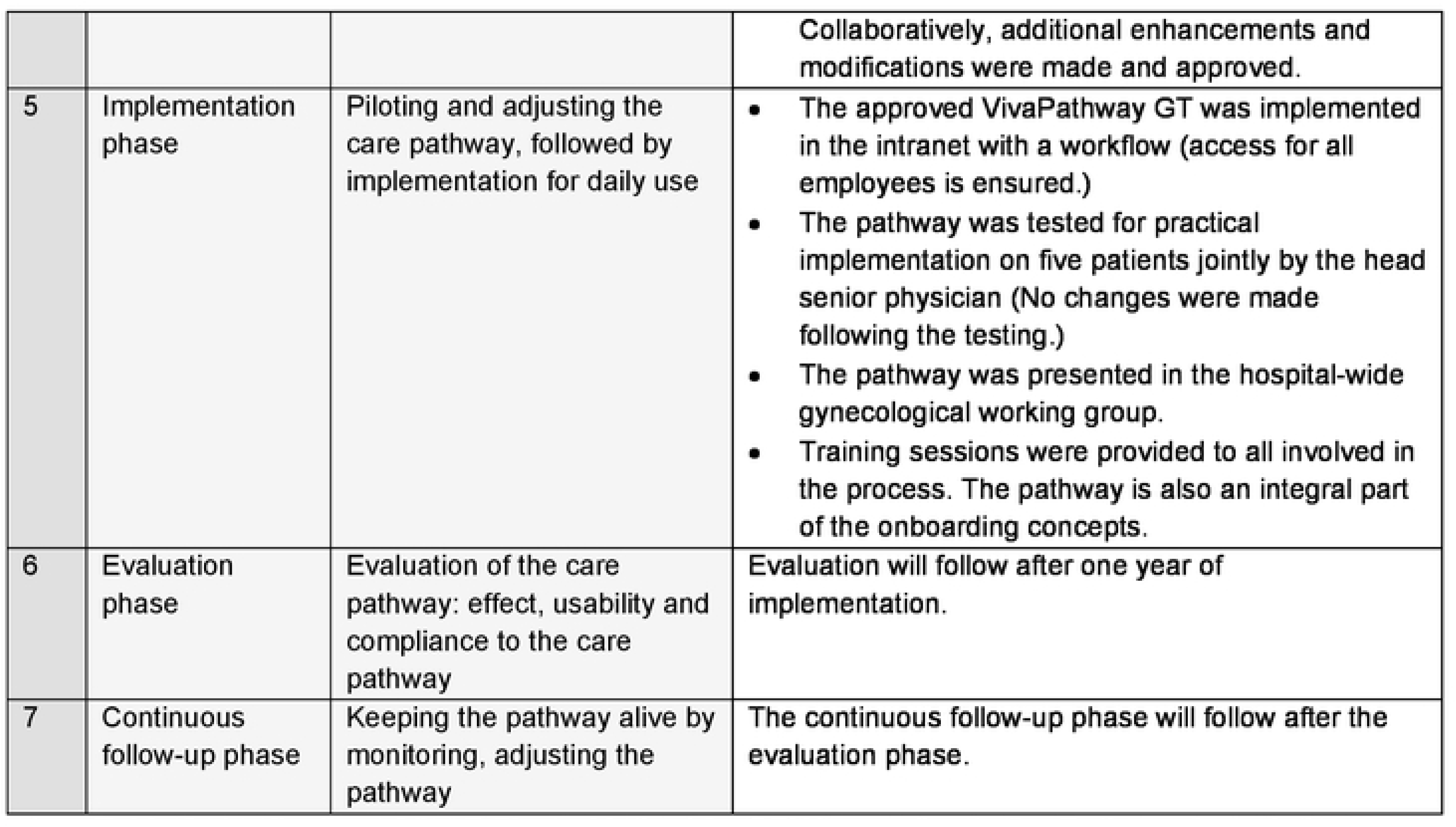
Application of the 7-Phase Method for CPW according to the Pathway Association.

| No. | Phase | Questions to consider | Interprofessional Approach for VivaPathway GT |
| --- | --- | --- | --- |
| 1 | Screening phase | Is a care pathway the right tool for the problem we need to solve; who will be the owner of the project / care pathway? | • The benefits of a clinical pathway for gynecological oncological patients were identified through research and legal requirements.<br>• Currently lacking an explicit pathway for gynecologic oncology.<br>• Until now there is an implicit pathway.<br>• Necessity to translate implicit standards into explicit pathways.<br>• Owner of the project is head senior physician of the gynecological department and a member of the hospital management. |
| 2 | Project management phase | Do we have the right people, time, budget to start a project; what are our goals? | • Right people: Certification criteria and other external requirements were assessed for stakeholders relevant to participating in the pathway.<br>• Time: A 2-year timeline was established, starting from the development of interview questions to the pathway's approval in the quality circle.<br>• Budget: An additional budget could not be considered.<br>• Goals: The aim is to transform the previously implicit pathway for patients diagnosed with C48.8 + 51 - C59 into an explicit pathway, considering all relevant external requirements. |
| 3 | Diagnostic and objectification phase | What is our current practice?<br>Based on four perspectives: own organization & team, vision of patient & family, available evidence & legislation, and external partners. | • The implicit pathway, the current practice, is examined through semi-structured interviews with all identified stakeholders. First with the leading department of gynecologists, then with all other involved professional groups.<br>• Interview results were investigated by Mayring analysis (Transcription and Correction, Formation of Analysis Section, Paraphrasing and Summarizing, formation of Categories/Codes, Application of Codes)<br>• In this way the organizations and team perspective were identified.<br>• Available research evidence was identified through guidelines. Legal issues were added.<br>• Vision of patient, family and external partners are missing and will be added after evaluation of one year. (Basis will be results of patient surveys and inquiries from referring healthcare providers.) |
| 4 | Development phase | Design and development of the care pathway, based on input from previous phases. | • Results of the Mayring analysis serve as the foundation for pathway development.<br>• Integration of insights from previous phases, including interviews, improvement suggestions, guidelines, inclusion of certification criteria and external requirements.<br>• Concluding discussion of individual steps in the quality circle, involving all relevant stakeholders. |
|  |  |  | Collaboratively, additional enhancements and modifications were made and approved. |
| 5 | Implementation phase | Piloting and adjusting the care pathway, followed by implementation for daily use | • The approved VivaPathway GT was implemented in the intranet with a workflow (access for all employees is ensured.)<br>• The pathway was tested for practical implementation on five patients jointly by the head senior physician (No changes were made following the testing.)<br>• The pathway was presented in the hospital-wide gynecological working group.<br>• Training sessions were provided to all involved in the process. The pathway is also an integral part of the onboarding concepts. |
| 6 | Evaluation phase | Evaluation of the care pathway: effect, usability and compliance to the care pathway | Evaluation will follow after one year of implementation. |
| 7 | Continuous follow-up phase | Keeping the pathway alive by monitoring, adjusting the pathway | The continuous follow-up phase will follow after the evaluation phase. |

**Flowchart 1:**
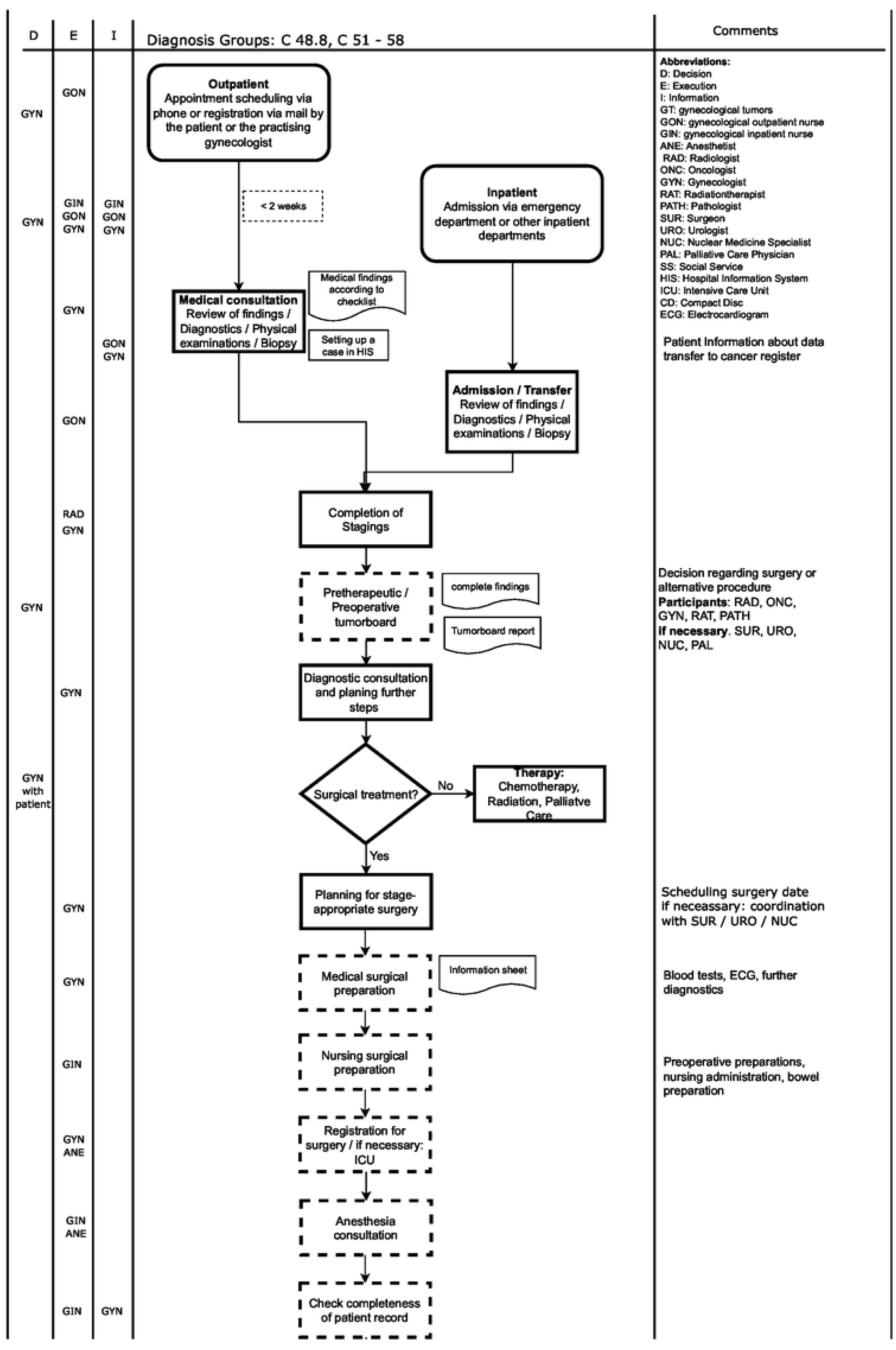
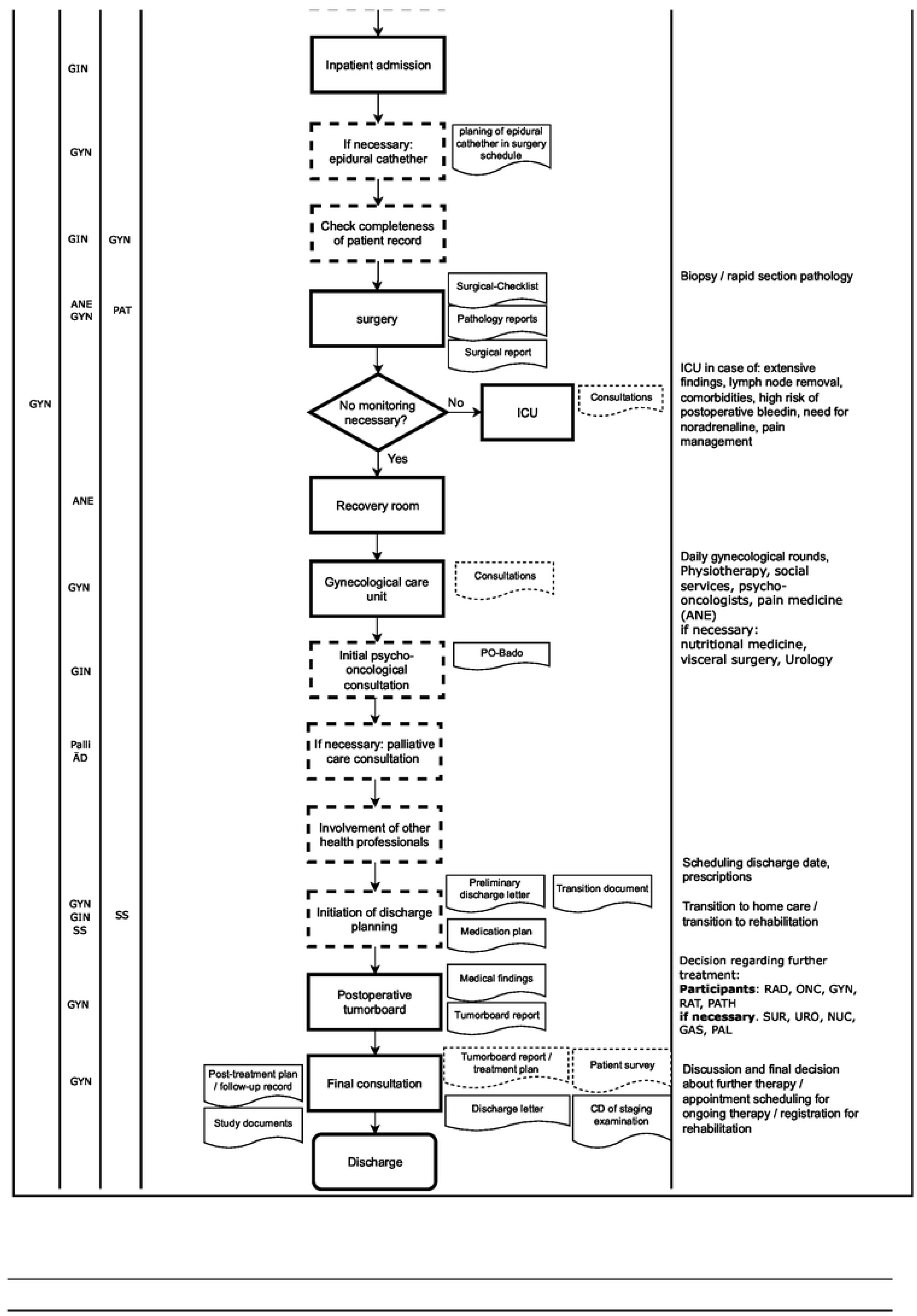
Clinical Pathway VivaPathway GT.

### 1. Application of the theoretical framework by the Pathway Association

The Results section demonstrates the application of the 7-phase method for clinical pathways to the VivaPathway GT. The first three columns illustrate the recommended approach by the Pathway Associations, specifying the phase’s name and the questions to consider during development of a clinical pathway. The meaningful outcome is evident in the last column, demonstrating the practical development of VivaPathway GT through an interprofessional approach.

### 2. The explicit VivaPathway GT

Next, the developed gynecologic oncological pathway VivaPathway GT is presented in a flowchart 1.

The analysis of gynecologists’ interviews indicates a notable consistency in their feedback regarding the steps in the care of gynecological oncology patients. The analysis reveals 14 categories/steps. Variations were observed in consultation requests with related departments. These were conducted through the hospital information system (KIS), telephone contact, and in-person meetings.

Moreover, all remaining stakeholders in gynecological oncological care were interviewed as well. They were asked regarding their involvement in the overall process.

Both, results of the gynecologists and all other stakeholder, formed the basis for discussions within the quality circle. During the interviews, all participants were also asked for improvement suggestions to enhance the future explicitly described process. These suggestions were incorporated into the interdisciplinary and interprofessional quality circle discussions.

Steps that were already included in the implicit pathway during the interviews are marked with a continuous line of the steps in flowchart 1. All steps, which were added during quality circle (on basis of the interview results), are marked with dashed lines. Consequently, it becomes evident that the implicit and explicit pathways primarily differ in that the explicit pathway is more comprehensive, containing more steps. The implicit pathway comprised 14 steps, and this was expanded with an additional 12 individual steps, resulting in a total of 26 steps.

### 3. Differences of implicit pathway to explicit VivaPathway GT

The main differences from the implicit to the explicit pathway can be summarized as follows:

- 14 implicit to 26 explicit steps
- Addition of the pre-therapeutic tumor board conference (certification requirement)
- More detailed preoperative preparation based on professional groups
- Documented checks for the completeness of patient records
- Active review of all possible post-op consultations by other professional groups (certification requirement)
- Initiation of discharge planning instead of general discharge (legal requirement)

## Discussion

This study illustrates the development of the gynecologic oncology clinical pathway in a tertiary hospital in Berlin within a participatory development approach. The outcome is based on the implicitly lived pathway and has been complemented with external requirements. The development process is mapped onto the theoretical framework of pathway association, allowing it to be easily transferred and applied by other physicians and managers on their initiatives. The tabular overview in table 2 provides a simplified method for transferring.

### Transition from Implicit to Explicit Pathways

The minimal deviations in the implicit pathway, as reported by the gynecologists, indicate that good coordination already exists without a written pathway. But when considering the pathway as a whole, it differs significantly from the pathway implicitly captured in the interviews. Though, the implicit pathway did not sufficiently anticipate the integration of other stakeholders.

### Top-down and bottom-up

Notable is the significant increase in the number of steps in the explicit clinical pathway, particularly highlighting the interprofessional approach in the care of gynecologic oncology patients. Köth et al. [15] already emphasized the necessity for all involved parties to be represented in the pathway. Through collaborative development based on interviews and a shared quality circle, this was ensured. Frequently mentioned by involved professional groups responsible for post-operative patient care was the desire for earlier involvement in the process. This includes physiotherapy, psycho-oncologist, nutritionist, social worker, etc., who are now all separately represented in the pathway. The hospital’s aim to become certified as an oncology center stems from leadership directives, representing a top-down approach to strategic planning.

The development of the clinical pathway VivaPathway GT is in line with the comprehensive findings in this field. Köth et al. [15] emphasized the importance of combining top-down and bottom-up strategies. Evans-Lacko et al. [16] underscore the essential role of involving both management and clinical staff in the development and implementation process. In addition, this approach also aligns with the success factors outlined by De Allegro et al. [17] in the implementation of clinical pathways, with a particular emphasis on the multidisciplinary aspect. In determining the clinical pathway, we integrate interdisciplinary and interprofessional elements, fostering collaboration among different stakeholders for a more holistic and effective approach. This fusion of top-down and bottom-up strategies, coupled with interdisciplinary collaboration, forms the foundation of the commitment to advancing health practices in gynecologic oncology on a scientifically informed basis.

Even when looking at other developments in healthcare, such as the development and implementation of Patient-Reported Outcome Measures (PROMs), similar success factors can be observed [18]. This underscores the consistent principles applicable across various healthcare interventions.

### Compensating for Missing IT-Infrastructure

The increase in steps from 12 to 26 can also be attributed to other reasons. Many checks have been introduced to ensure that previous steps are carried out and documented. Similarly, several steps of different professional groups have been introduced in the preparation for surgery. Both could theoretically be supported by an IT infrastructure, as well as the post-operative consultation of other professional groups. In this case, significantly fewer steps would be necessary. For example, a hospital information system (HIS) could automatically request a consultation with a physiotherapist once a specific diagnosis and procedure have been documented. Since this functionality is not currently available in the hospital’s HIS, it has been separately integrated into the explicit clinical pathway.

A comparison can be drawn between the development and implementation of clinical pathways and another health intervention, such as Patient-Reported Outcome Measures (PROMs). The study by the Bertelsmann Foundation [18] described six success factors needed for the successful implementation of this intervention. Therefore, a relevant success factor is supportive IT. This would even allow for the inclusion of PROMs assessments within the IT-based clinical pathway.

A competent IT infrastructure supporting the clinical pathway serves as an essential prerequisite for comprehensive documentation of patient records. It is through careful documentation that a meaningful evaluation can be conducted in the Plan-Do-Check-Act (PDCA) cycle. Another argument for the relevance of the IT-based representation is provided by Askari et al. [19]. He found, in a systematic review, an increased adherence to clinical pathways when software is used to support the pathway. Also, Ronellenfitsch [20] concludes that integration of clinical pathways into hospital information systems in the course of digitization in healthcare could be beneficial for the application and acceptance. Not only integration into HIS, but also the development of an innovative process mining solution is emphasized by Munoz-Gama et al. [21]. He explicitly calls for close collaboration between health professionals, IT specialists, and business process managers to develop a supportive evidence-based system.

### Underutilization of Clinical Pathways

For the underutilization of clinical pathways in healthcare, there are various reasons. In this case, it did not result from health professionals’ unwillingness to adhere to standards. The results indicate that gynecologists and all other stakeholders wish to work according to guidelines and internal standards. The gynecologists have already been working based on an implicit pathway, and all other stakeholders also adhere to their standards. However, these have not been consolidated for the care of gynecologic oncology patients until now.

However, limited financial and personnel resources Köth et al. [15] pointed out the necessity of effective outcomes required for the development and implementation of pathways, given the substantial resource investment. This assumption aligns with the results of the Bertelsmann Foundation [18] regarding the introduction of PROMs. Financial support is described as one of the success factors for implementation alongside other forms of support.

Since 2017, quality contracts in Germany have offered opportunities to allocate additional resources in clinical settings, for example for pathway developments [22]. They aim to provide additional incentives for improved care. Explicit pathways are a necessary prerequisite for a quality contract and its evaluation. However, currently, this is offered for eight medical conditions such as endoprosthetics or obstetrics. In future, this could potentially be utilized as financial support to develop and implement pathways in hospitals with a participatory approach.

### Addressing the research question and validating the hypotheses

Considering all results, the research question can be answered and the hypotheses can be confirmed:

#### Research Question

*How can an implicit pathway in gynecologic oncology be translated into an evidence-based, explicit care pathway in a tertiary care hospital in Berlin?*

Answer: The research question is addressed through a comprehensive process involving stakeholder interviews, analysis, and the development of the VivaPathway GT. The translation of the implicit pathway is achieved by integrating insights from interviews with health professionals and incorporating external requirements.

#### Hypothesis 1

The interprofessional translation of implicit pathways, guided by stakeholder interviews, facilitates the development of explicit pathways.

Confirmation: The stakeholder interviews and subsequent Mayring analysis form the foundation for the explicit VivaPathway GT, demonstrating that an interprofessional approach is effective in translating implicit pathways.

#### Hypothesis 2

Integrating external requirements, especially legal mandates, during development establishes a vital connection between external demands and internal structures, resulting in a holistic and evidence-based pathway.

Confirmation: External requirements, including legal mandates, are integrated into the VivaPathway GT during the development phase. This ensures a connection between external demands and internal structures, aligning the pathway with evidence-based practices.

The successful development of the explicit pathway, VivaPathway GT, validates both hypotheses, highlighting the importance of stakeholder collaboration and the incorporation of external requirements in achieving a comprehensive and evidence-based care pathway in gynecologic oncology.

## Conclusions

In conclusion, this study presents a valuable contribution to the development of clinical pathways. The results offer a helpful guide for other healthcare professionals facing similar challenges and requirements. However, it is important to acknowledge that disruptions may occur in different settings, and while the health professionals in this study were willing to implement a standardized pathway, it remains a sincere resource challenge due to the significant time and effort required for a participatory approach involving all stakeholders.

The necessity of transitioning from implicit to explicit pathways cannot be overstated, as demonstrated by the formalization, standardization, and documentation of clinical workflows. The study aligns with previous research underscores the significance of multidisciplinary, both top-down and bottom-up approaches in this process in order to adequately consider the needs of all stakeholders.

Furthermore, the study highlights the crucial role of IT support in securing, refining, and evaluating pathway processes, with the potential to incorporate patient perspectives and adapt to evolving structures and external mandates over time. However, the explicit description of a pathway is a precondition for its integration into HIS and process mining tool.

In the face of financial and resource limitations, initiatives such as quality contracts offer opportunities to allocate resources for pathway development and evaluation, ultimately contributing to improved patient care.

In addressing the research question and validating the hypotheses, the study demonstrates the effectiveness of an interprofessional approach guided by stakeholder input and the integration of external requirements in developing evidence-based and comprehensive clinical pathways. The successful development of the explicit pathway, VivaPathway GT, underscores the importance of stakeholder collaboration and the incorporation of external mandates in achieving a robust and adaptable care pathway for gynecologic oncology patients. However, ongoing evaluation and adaptation are essential to ensure the pathway’s effectiveness and relevance among changing healthcare landscapes.

## Data Availability

No, the data are available from the author and are not publicly accessible. The reason for this is due to the methodology of the study: in the interviews, we always chose only one profession per interview participant (except for the gynecologists). Making the data publicly available would not maintain the promised anonymity of the participants, which would be in conflict with the ethical guidelines of our research. For this reason, we have decided to make the data available only upon direct request, in order to protect the privacy and anonymity of the interview participants. Data are available from the first author for researchers who meet the criteria for access to confidential data.

## Authors contribution

Anne Büscher

- Conceptualization
- Formal Analysis
- Investigation
- Methodology
- Project Administration
- Writing – Original Draft Preparation
- Writing – Review & Editing Nicolle Reinhold
- Conceptualization
- Validation Joachim Kugler
- Conceptualization
- Supervision

## Supporting information captions

## Competing Interests Statement

The authors of this manuscript declare the following potential competing interests in accordance with PLOS guidelines:

**Anne Büscher:** As the administrative manager at Vivantes Klinikum Friedrichshain, I am administratively involved in the implementation and management of the clinical pathway discussed in this manuscript. My role involves general administrative and managerial functions within the whole hospital.

**Dr. Nicolle Reinhold:** Dr. Reinhold is a senior physician in gynecology at Vivantes Klinikum Friedrichshain and directly involved in patient care and the development of gynecologic oncology practices.

All interviews conducted for this research involved participants associated with Vivantes Klinikum Friedrichshain, and their identities are familiar to the authors due to their roles within the hospital. No financial interests, employment, consultancy, or any other competing interests that could compromise the objectivity of this research are present.

This statement is made to ensure transparency and adhere to PLOS policy on competing interests.

